# Differences in power-law growth over time and indicators of COVID-19 pandemic progression worldwide

**DOI:** 10.1101/2020.03.31.20048827

**Authors:** Jack Merrin

## Abstract

An automated statistical and error analysis of 45 countries or regions with more than 1000 cases of COVID-19 as of March 28, 2020, has been performed. This study reveals differences in the rate of disease spreading rate over time in different countries. This survey observes that most countries undergo a beginning exponential growth phase, which transitions into a power-law phase, as recently suggested by Ziff and Ziff. Tracking indicators of growth, such as the power-law exponent, are a good indication of the relative danger different countries are in and show when social measures are effective towards slowing the spread. The data compiled here are usefully synthesizing a global picture, identifying country to country variation in spreading, and identifying countries most at risk. This analysis may factor into how best to track the effectiveness of social distancing policies and quarantines in real-time as data is updated each day.

## 2 Introduction

Mathematics is essential to predict and control the course of a pandemic. One approach is mathematical modeling [1]. Many epidemiological models derive from the SIR (susceptible, infected, recovered) model [2] or some more sophisticated versions of it. If there are susceptible, infected, and recovered (or removed individuals), then there are standard shaped curves and predictions you can make based on differential equations, provided you exactly know the transmission rate and recovery rate.

These parameters may only be possible to determine from a retrospective analysis. One parameter is *R*_0_, the basic reproduction number, which represents the number of people an infected person typically can infect over their progression. Clearly, if *R*_0_ is much larger than 1, then there is a significant initial exponential growth of cases that sweep through most of the population. China and South Korea have mostly blocked the spread of COVID-19 by enacting social policies and testing so *R*_0_ can definitely be reduced or modulated. The SIR equations are also entirely deterministic, while in real systems, there is some stochastic element of variation. SIR equations do not have an analytical solution so they are more difficult to solve, and determining the fitting parameters requires sophisticated statistics. Usually for an epidemic like the flu everything stays open so the parameters are more or less the same over the entire trajectory of the analysis. Things start to become very complicated if different countries enact different social policies over time with different degrees of strengths. SIR model parameter themselves would then become functions of time complicating the analysis. Spreading the virus may also depend on differences in geography, social networks, social dynamics, or individual behaviors.

Another approach is not to have any mechanistic mathematical model at all, but to use methods of error analysis [5,6,7,8]. One can fit the data to different phenomenological functions observed in the past, such as what happened in China over the last several months to make comparisons.

In this study, we attempt to analyze differences in the spread of COVID-19 in many countries where growth is rapid to 1000 cases or more as of March 28, 2020. We use the least-squares curve fitting of power-laws and introduce the danger indicator to compare the rate of spread in various countries. Both indicators reveal similar information in the current stage of the pandemic. The data analysis follows the proposal of Ziff and Ziff [3], where an early stage involves exponential growth, and a middle stage exists where there is power-law growth. The power-law exponents allow us to make reliable short term predictions of a few days of the worst-case scenarios in hot spot countries compared to that of the usual assumption of exponential growth. Timescales for mitigation may be three weeks or more and can be tracked by this analysis.

These predictions and indicators may be relevant for estimating the burden on medical infrastructure or influence policy decision making before countries enter mitigation or rather the virus completely sweeps through. Of all countries so far, only China and South Korea have achieved mitigation compared to other countries where there is still significant power-law growth. This analysis identifies some countries which are in a highly critical situation with expectations of more positive cases and deaths than occurred in China, while others have quite some time to achieve a resolution.

## 3 Methods

### 3.1 log-log plots

In this paper, we display the number of measured positive cases (data set WHO [9]) as a function of time on plots where the *x* and *y* variables are both logarithmically scaled. This is referred to as a log-log plot. We will try to distinguish between exponential growth and power law growth. Here we show exponential growth on a log-log plot appears like an exponential curve on a linear plot.

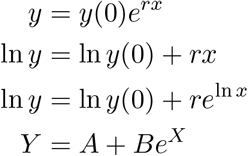

Here we show power law growth on a log-log plot appears linear.

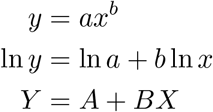

Increasing power law functions on a log-log plot have positive slope. There can be a high degree of overlap with the power law growth and exponential growth in the middle region, but the long term behaviours are different as shown in figure 1.

**Figure 1:**
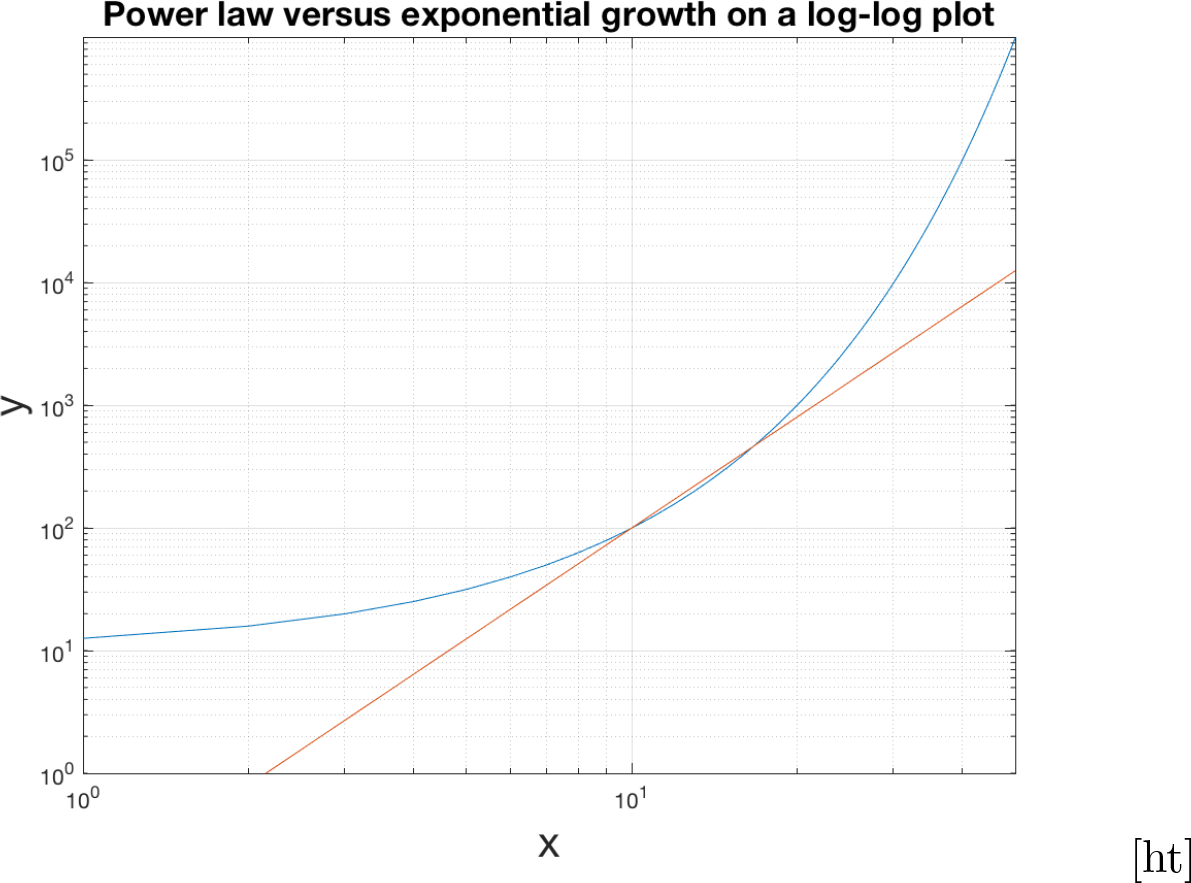
Here we plot 0.1*x*^3^ vs *x* in red and 10*e*^0.23*x*^ in blue.

**Figure 2:**
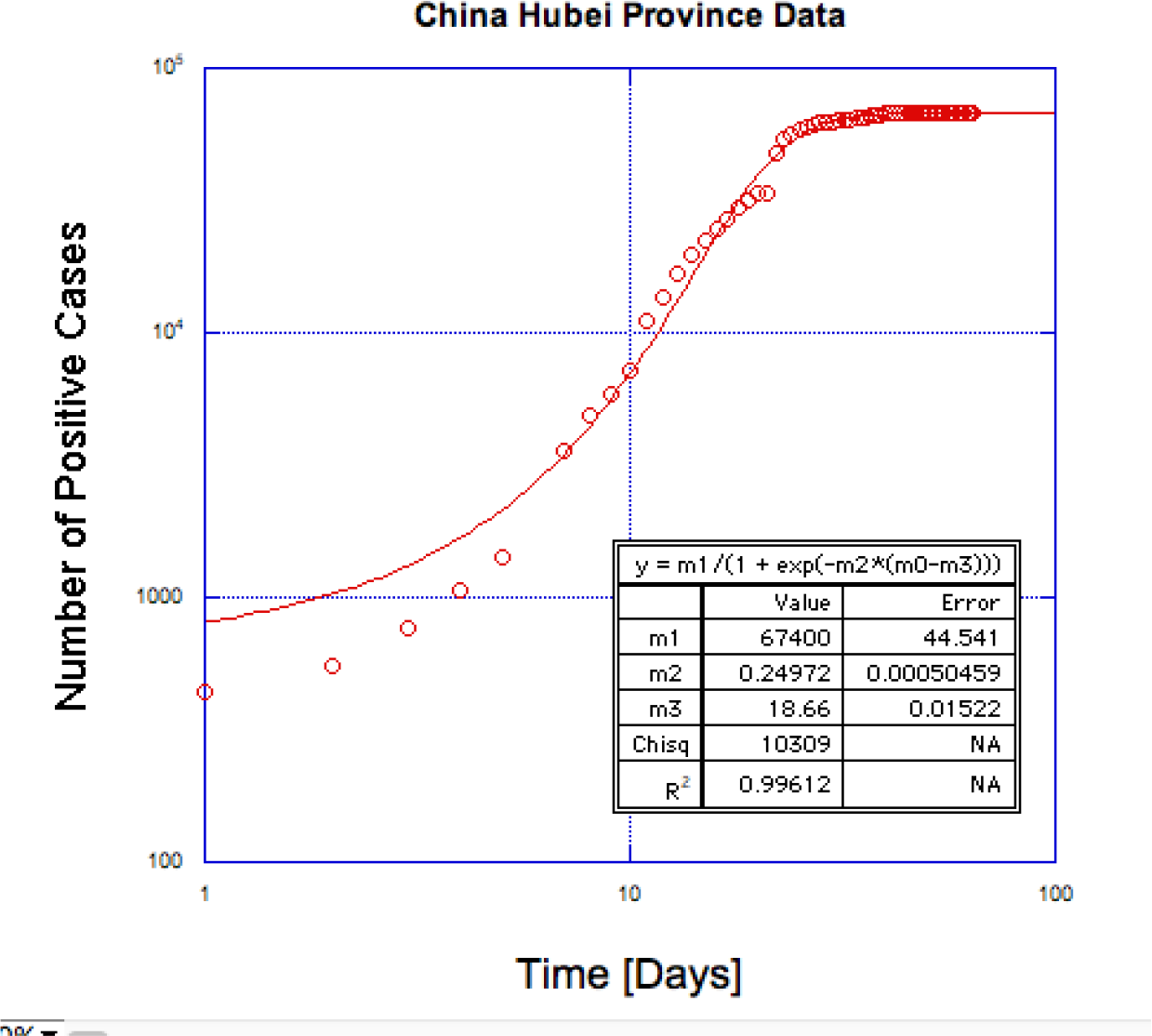
Hubei China data [9] as a basic model fit to logistic curve with Kaleiagraph. Using the logistic curve fit we can get some idea of how beta progresses over the course of the entire model sequence of events using the analysis in this paper. This is done because the actual data is slightly corrupted.

### 3.2 Linear Least Squares Curve Fitting

In this paper, we use error analysis [5,6,7,8] and curve fitting to analyse the progression of positive cases. In weighted least squares curve fitting, one finds the parameters which minimise the chi squared function.

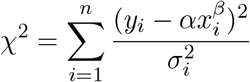

As we saw before, a power-law function can be transformed into a linear function on a log-log graph. There is always an analytical solution to the curve fit of a straight line and thus we can determine *α, β*, and their uncertainties quite objectively in a standard and simple way. The transformation is

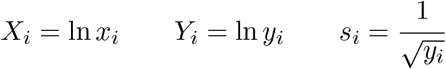

with *A* = ln *α* and *B* = *β*. Then,

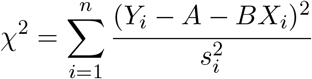

### 3.3 Fitting a straight line

If we want to fit data to a straight line, then we need the coordinates of the statistics *x*_*i*_, the measured values

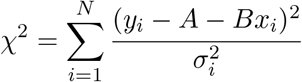

*y*_*i*_, and the uncertainties in the values *σ*_*i*_. Then we have for chi-squared.

In our cases, *x*_*i*_ is the number of days of the progression of the disease since 100 cases. *y*_*i*_ is the reported positive tested cases at *x*_*i*_. You cannot repeat the measurement, so we have to estimate the typical uncertainty. We have to use the square root *N* rule. For example, if there are 100 cases, then the uncertainty would be expected to be about 10 cases with a relative uncertainty of 10 percent. If there are 10,000 cases, then the uncertainty would be expected to be about 100 cases with a relative uncertainty of 1 percent. The relative uncertainty is given by

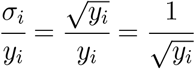

More recent points should be weighted more strongly because their relative error is less. A more exact curve fit incorporates the use of error bars.

To solve for *A* and *B* you calculate 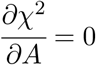 and 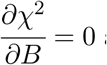 and solve the resulting system of linear equations. We use the notation,

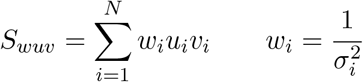

The uncertainties in the parameters can also be directly computed using propagation of error.

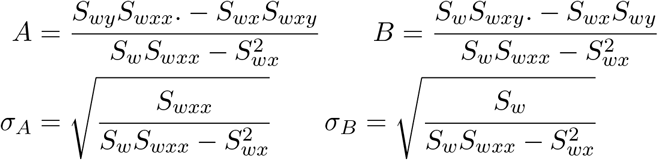

### 3.4 Propagation of error

If a statistic and its uncertainty are known as *x ± σ*_*x*_ then we can transform *x* into *f* (*x*) and the new uncertainty is given by propagation of error.

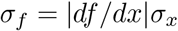

If *A* = ln *α ± σ*_*A*_ then *σ*_*α*_ = *e*^*A*^*σ*_*A*_. Of course, *β* = *B* and the uncertainty is obtained directly from the linear curve fit. We also used the fact that *Y* = ln *y*_*i*_ so

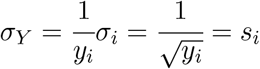

### 3.5 Other methods of determining beta

It was also attempted to do an unweighted fit with the following formulas [10], but there is not much difference in beta for the two methods. See the table at the end of the results. The weighted fit is preferable to compute the error bar in beta.

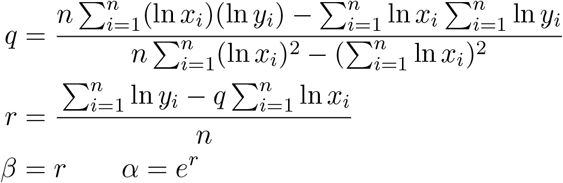

In the future, it may also be attempted to perform a maximum likelihood estimation of the power law exponent since there are some caveats to the weighted method presented here, such as assuming no uncertainty in the × coordinate [11]. Another alternative is to use a software like Kaleidagraph, which does a nonlinear least squares analysis based on the Levenberg-Marquardt method. Using a software package like that is cumbersome to deal with all the datasets for each time window as described in the next subsection. Also since you didn’t write the program, it is not clear that you understand what was computed is right. The weighted linear least squares method is considered sufficient for the time being and works very well automatically in Matlab scripts and produces the uncertainty in beta.

### 3.6 Moving window procedure

One last detail is that the data must be linear on a log-log plot, or the points that don’t match are outlying. Outliers skew the values of *α* and *β* and hinder any attempts to make accurate predictions. The initial phase of the data is not power-law, so we restrict ourselves to measuring the last 7 days as a measure of progress in a country. A moving window has the added benefit that you can have a moving window of seven days to see the progress over time as we move the windows backward from now. If the curve stays in a power-law regime, the plot of the exponent is flat versus time. That would be the latest overall exponent. It will also reduce in magnitude as the rate of infections decreases. The data from China and South Korea follow this pattern and do not fit a power law for all times because an end-stage of no new infections occurred. Basically, a moving window is similar to quantifying the magnitude of slope on a log-log curve over time as an indicator of disease progression.

Only countries that had reached 1000 cases or more were considered as hot spots for analysis. There was a lower cutoff of 100 cases where the data is often irregular. Times before 100 cases are less relevant, and you can’t even plot time zero on a log-log plot because log 0 is undefined. More cases bring more predictability. The moving window procedure also has the benefit that you don’t have to manually and arbitrarily pick when the power law regime starts in the analysis. You can just track it and see if it locks in for the most recent data. Some countries go power law after 1000 cases or more but the plots presented in the results start at 100 cases to get a better picture.

### 3.7 Measure of Danger

In principle, if you limit the spread of the virus as in South Korea and China, then the power-law growth is reduced to a zero exponent. A measure of danger is the time it would take to infect the remaining population of a country versus the time required for a useful quarantine to limit growth. The indicator *D* we introduce here does just that.

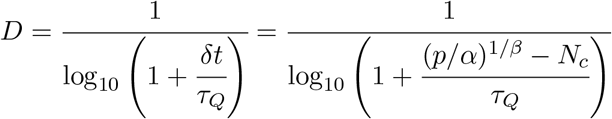

The *D* indicator depends on several variables.

- *p* is the population of the country.
- *N*_*c*_ is ideally the current total number of infections, but we only have measured positive cases.
- *α, β* are the fit parameters to the number of positive cases at the current time,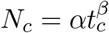.
- *τ*_*Q*_ = 14 days is the quarantine time.
- *δt* is defined as the time it takes for the power-law to extrapolate to the total population of that particular country.

The *D* indicator is a positive function with the following properties. When *δt* ≫*τ*_*Q*_ then *D →*0. The country would have much time to implement a social distancing strategy. When *δt* ≪*τ*_*Q*_ then *D* ≫1. If *D* is large, and more rapid social distancing is more crucial to reduce future infections. Since most countries will not exhaust their populations for some time from March 28, 2020, then *D* as a function of time looks very similar to just *β* as a function of time, so I choose not to plot it or report the values. If significant infection occurs relative to population size, we should probably use measures of spread more in lines with *D*.

### 3.8 What we are looking for with this method

People are interested to know when the number of new cases saturates. And if they do saturate, is this substantially lower than the total population, or does it sweep through. South Korea and China datasets are most interesting because they show it can stop before sweeping through everyone (67400 cases Hubei China). The Hubei province data is the most interesting because it shows an entire trajectory of the infection. There is one problem with China Hubei province data. The criteria for a positive test was extended to include lung imaging on February 13. Discontinuities from differences in testing recording are bad for a moving window analysis that we use. Instead, as an example, we make an interpolating function, which is a logistic curve. We can see what the moving window method predicts as a basic model on ideal data where testing would have been perfect. For all other analysis in this paper, we assume no interpolating function. Also the other China datasets with better measurements for different provinces look like the same progression of beta as we show in figure 3 for this toy model. In summary, we should keep in mind the dropping of beta to zero is what we want to happen as the outbreak comes under control in different countries. If beta is increasing that indicates things are getting worse.

**Figure 3:**
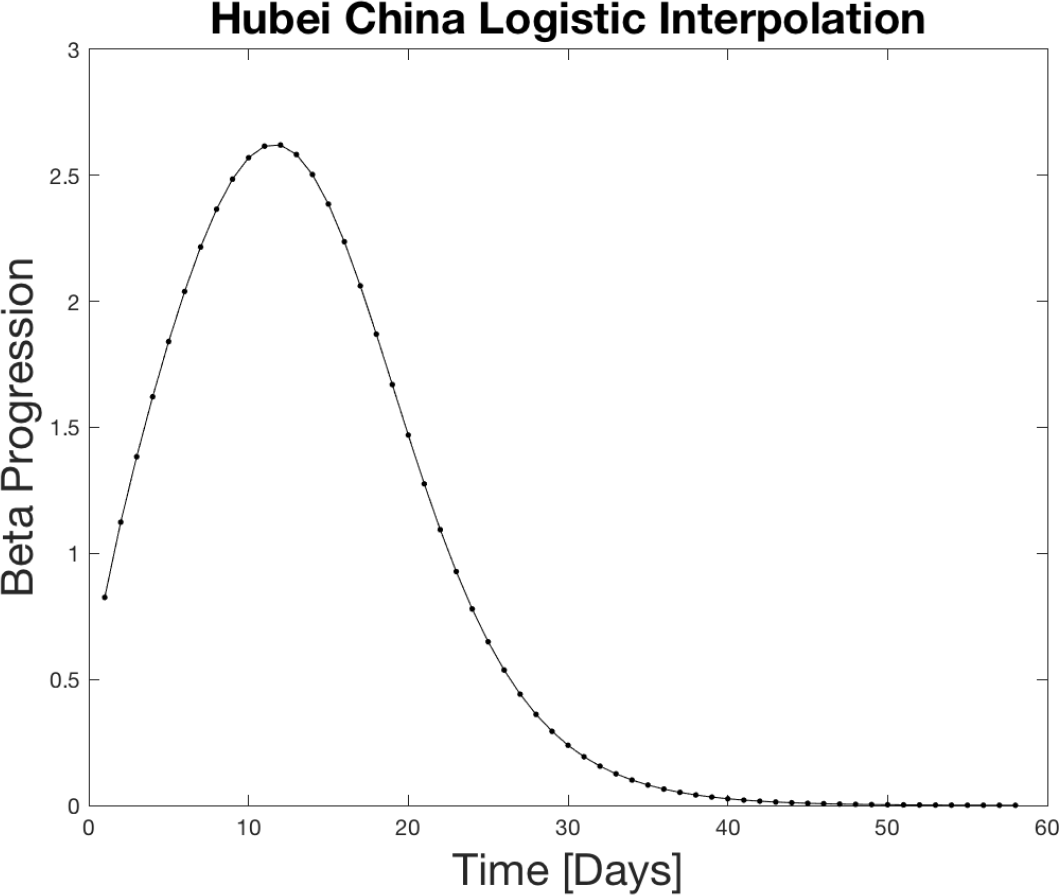
7 day moving window analys of the power law exponent for the Hubei China curve fit function. It starts low, reaches a peak around 2.5, then flattens out back to zero as the condition becomes under control. Matlab analysis of the interpolating function from figure 2.

**Figure 4:**
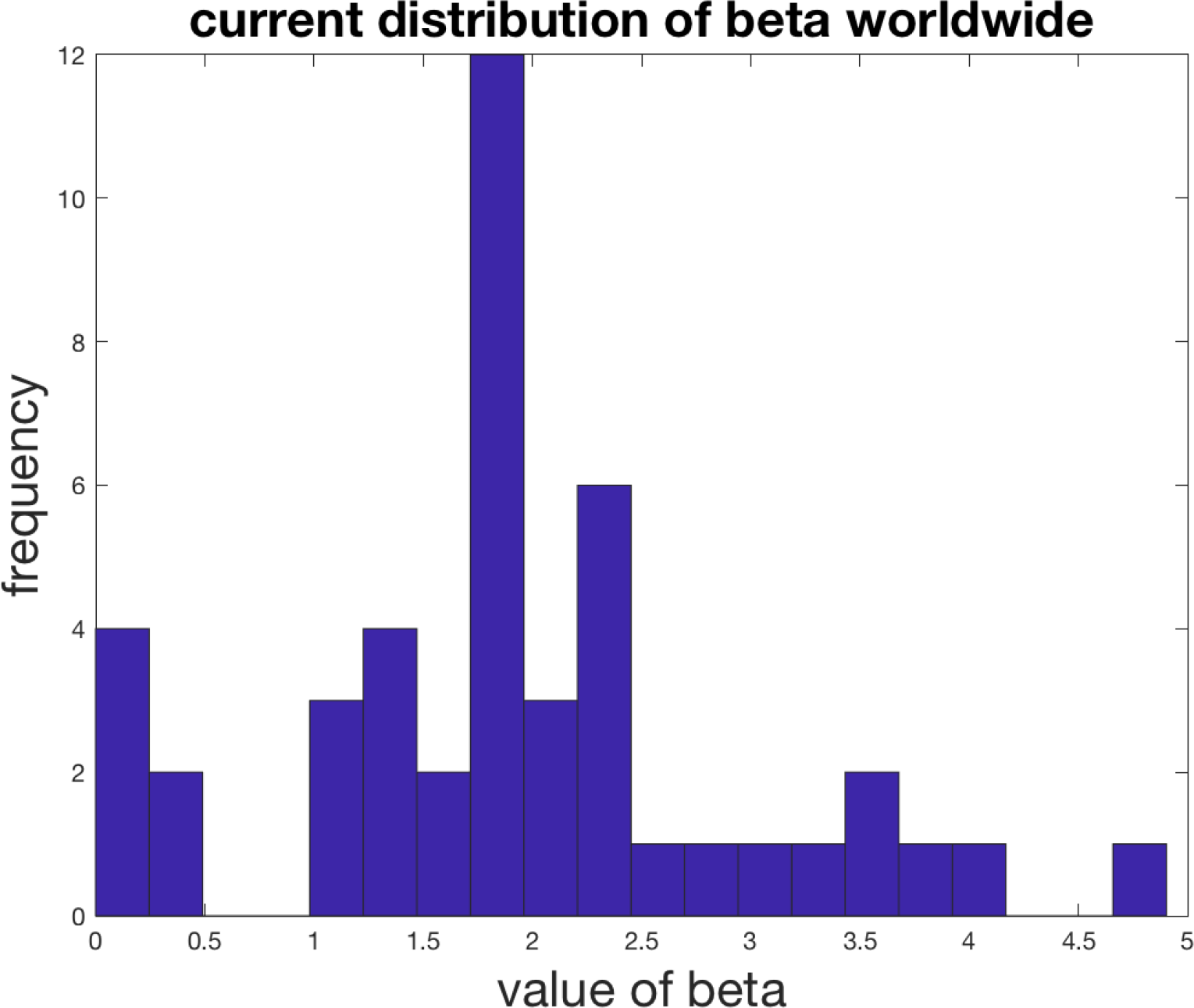
Beta correlates with how drastic the situation is with different countries currently. Beta less than 0.5 can be considered as under control, Beta greater than 3 indicates a potentially dangerous situation.

## 4 Results

### 4.1 Power law fits for positive cases

The black dots represent the actual data. The curve fit for the last seven days is extrapolated as the black line. The top edge of the graphs is set to the population of the various countries. The x-axis spans 300 days. One indication of the severity of the spread is the relative slope of the power-law on the log-log plot. A higher slope means things are increasing faster soon. Notice for the case of the US that the projected time scale is less than 100 days if the curve does not flatten out. The extrapolation has relevance for setting social policy as a worst case scenario.

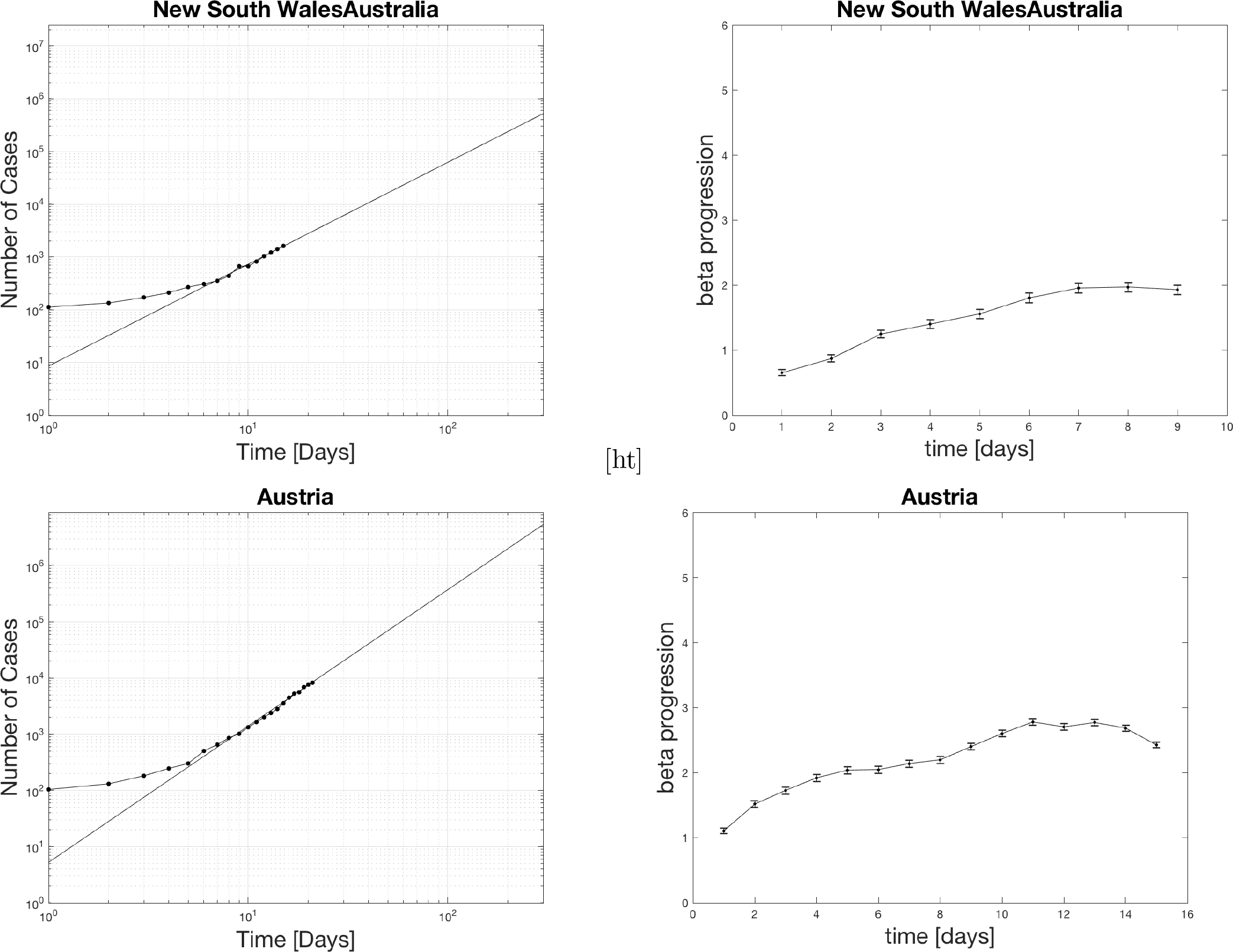

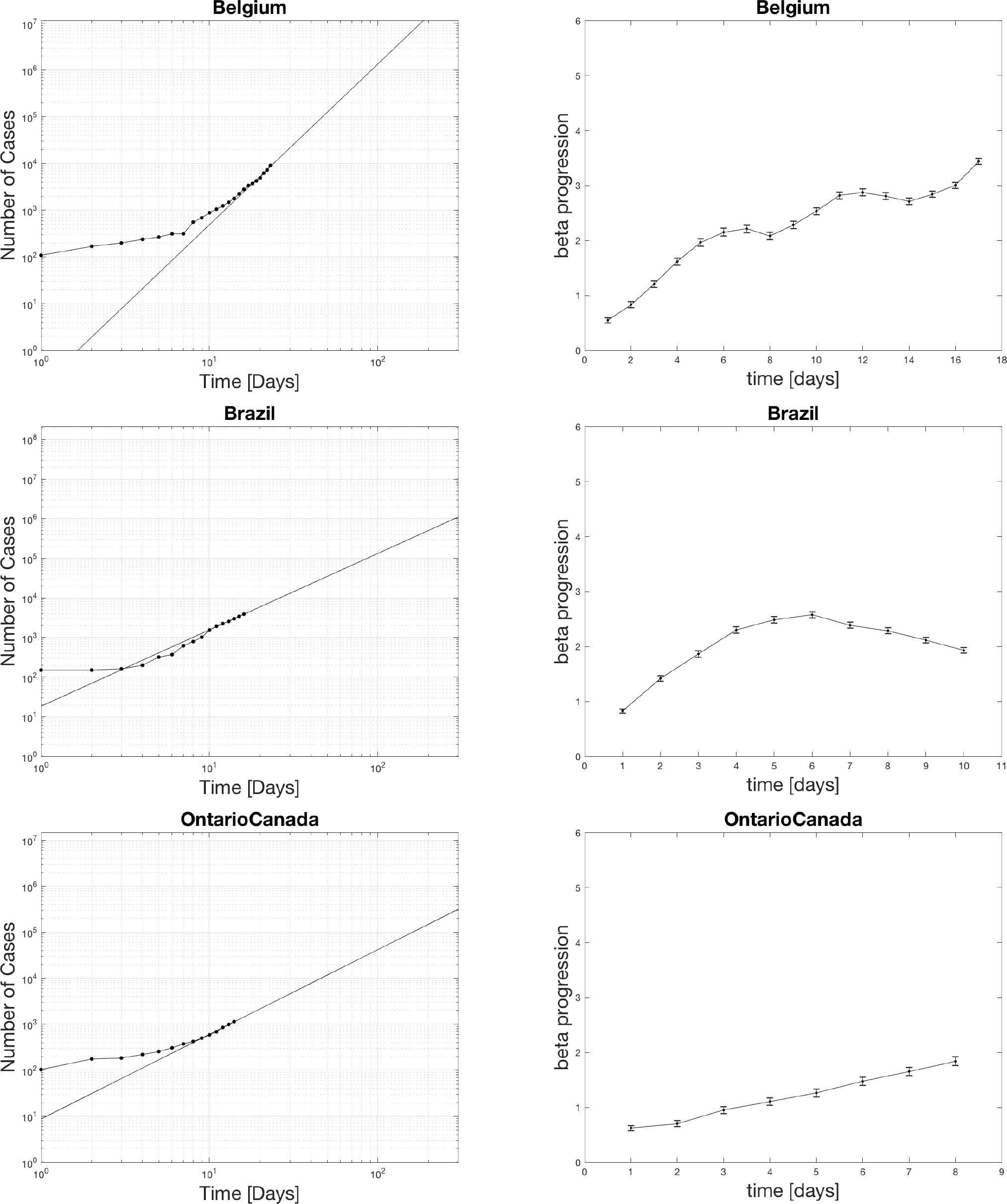

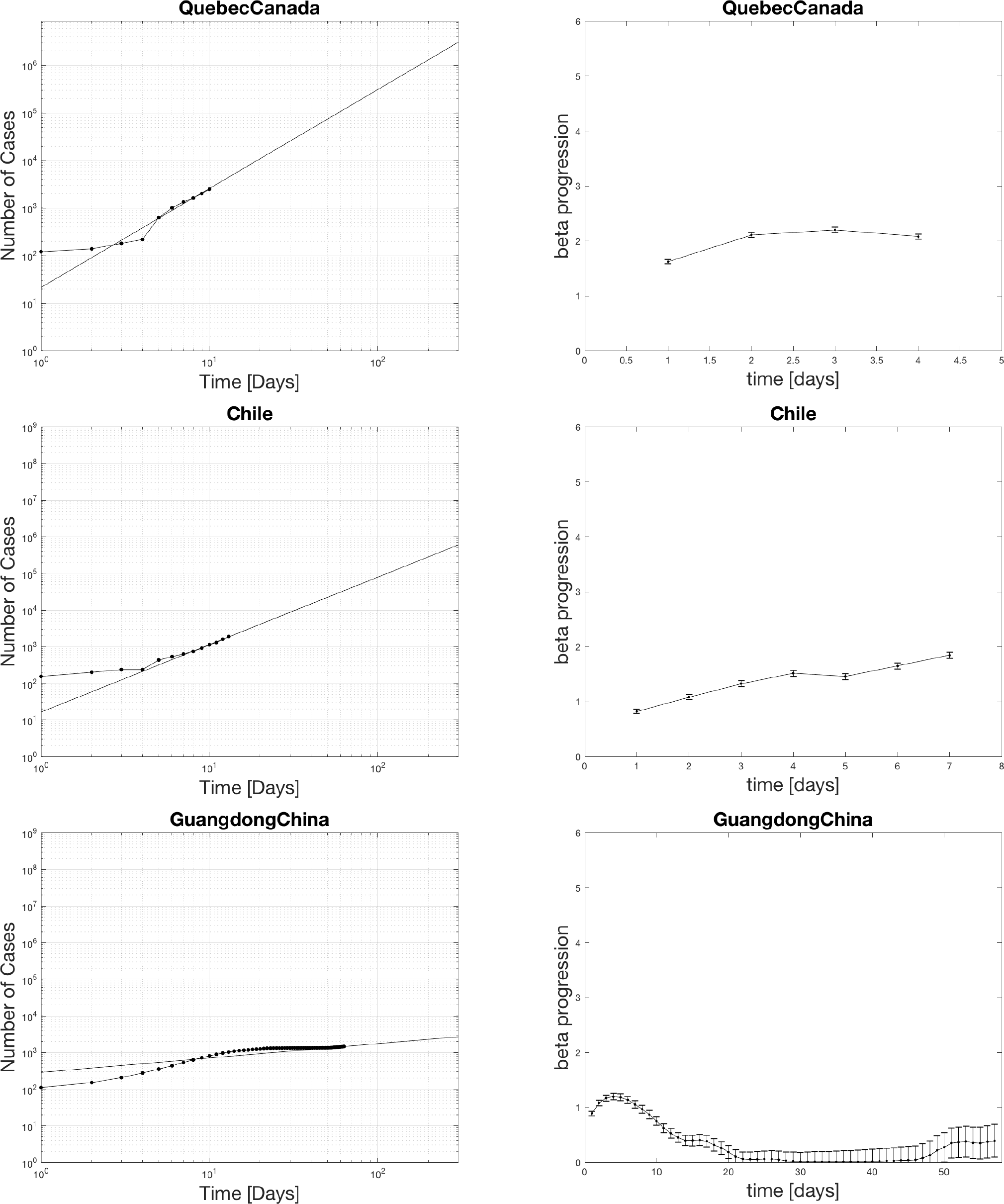

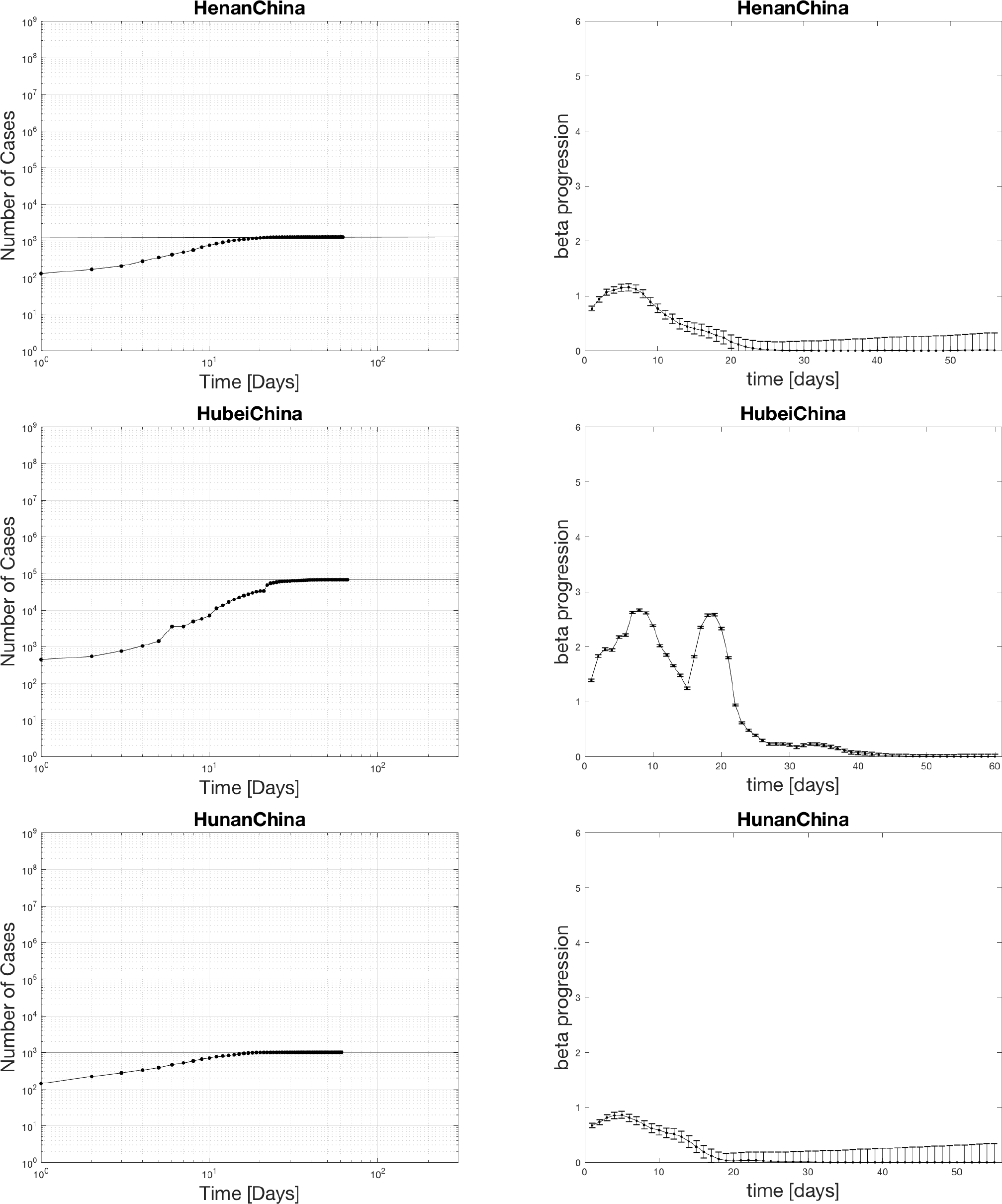

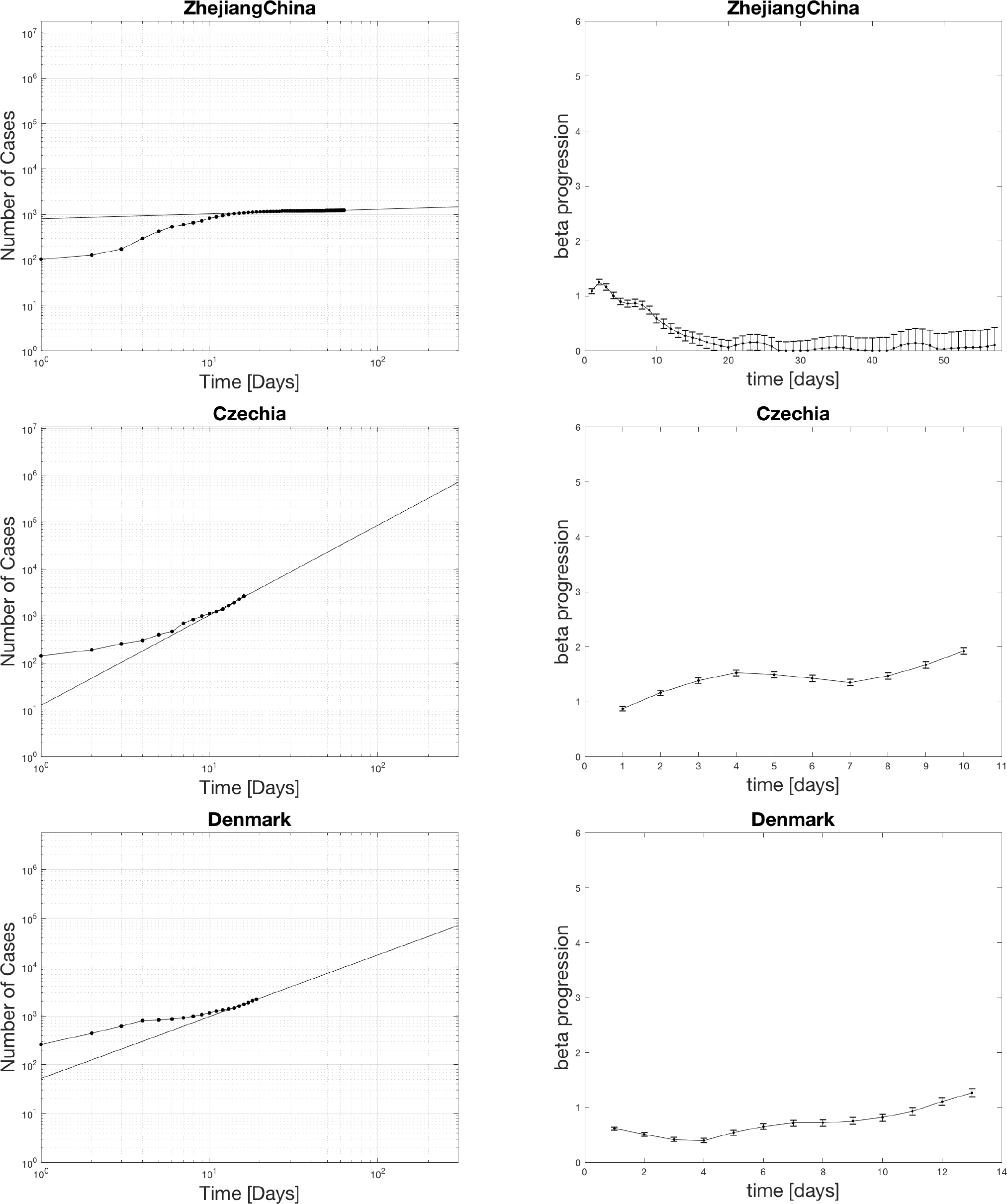

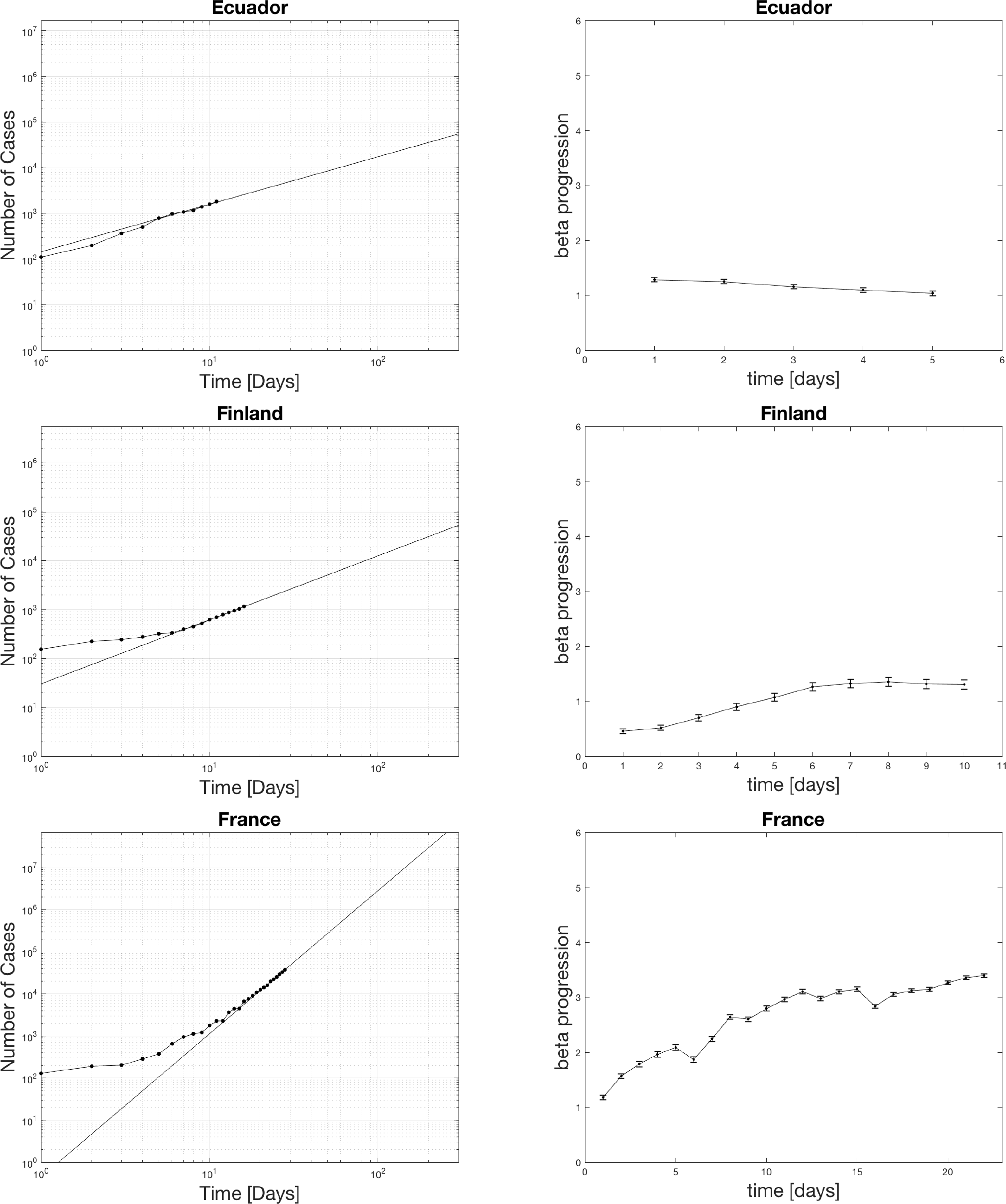

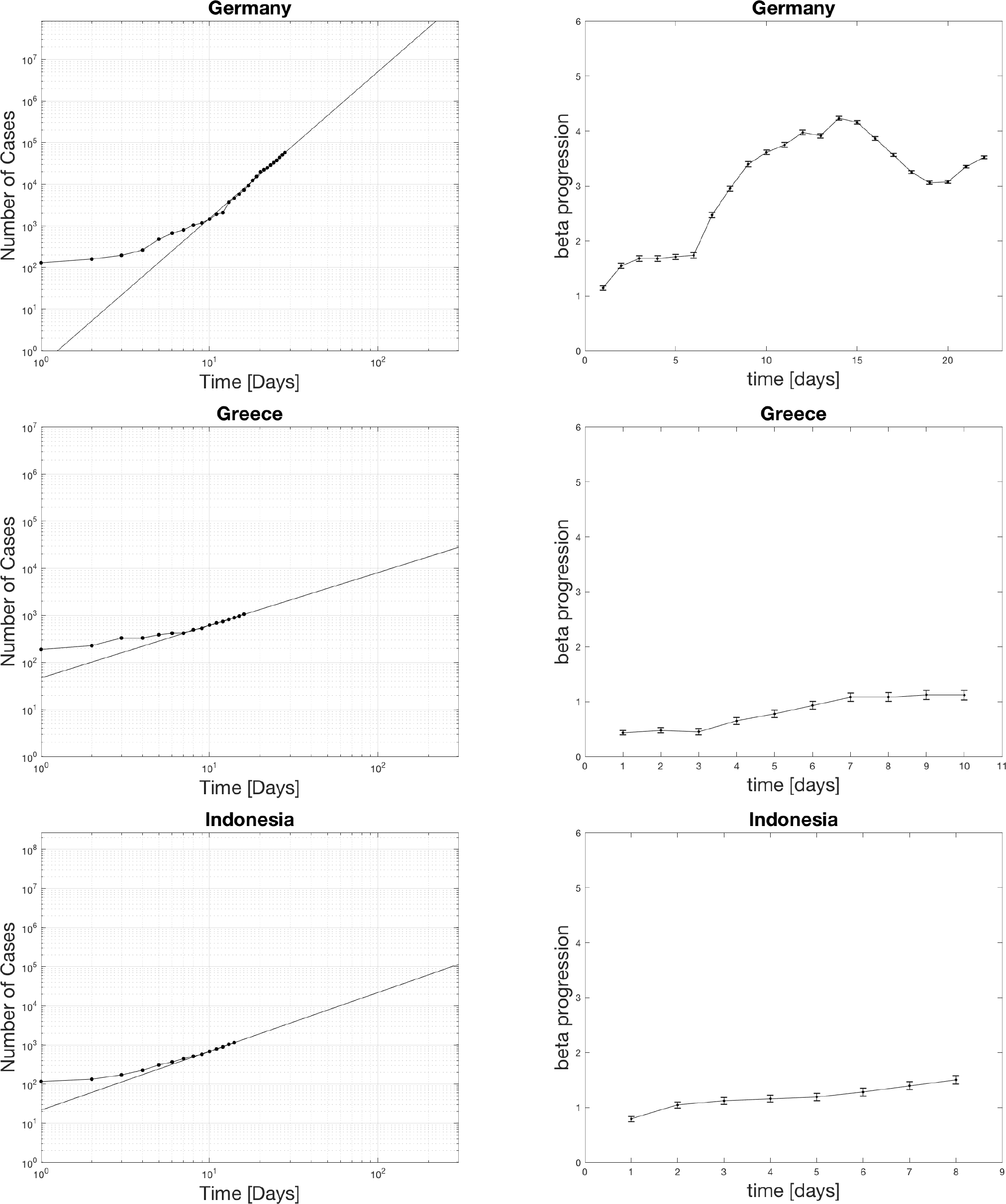

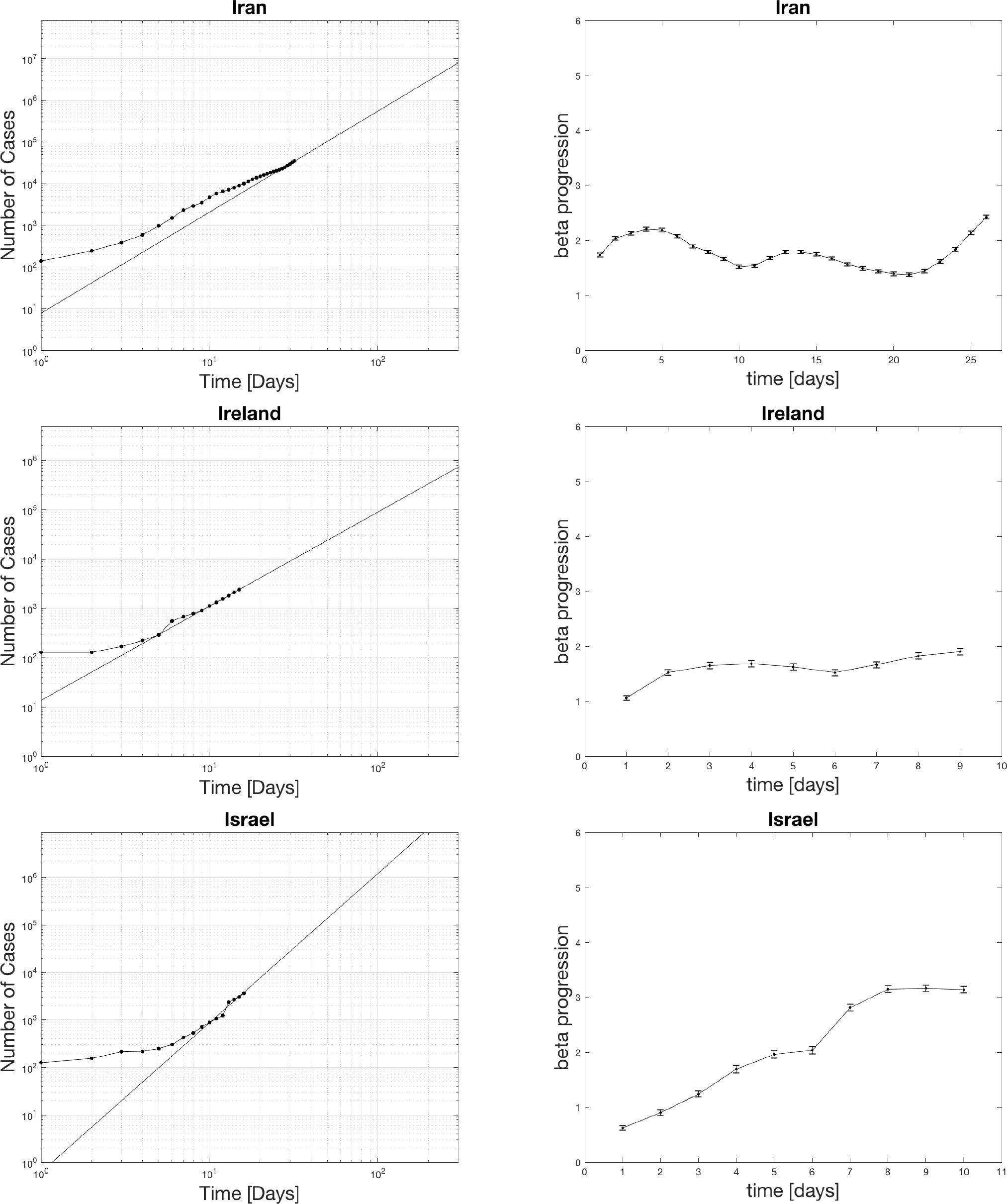

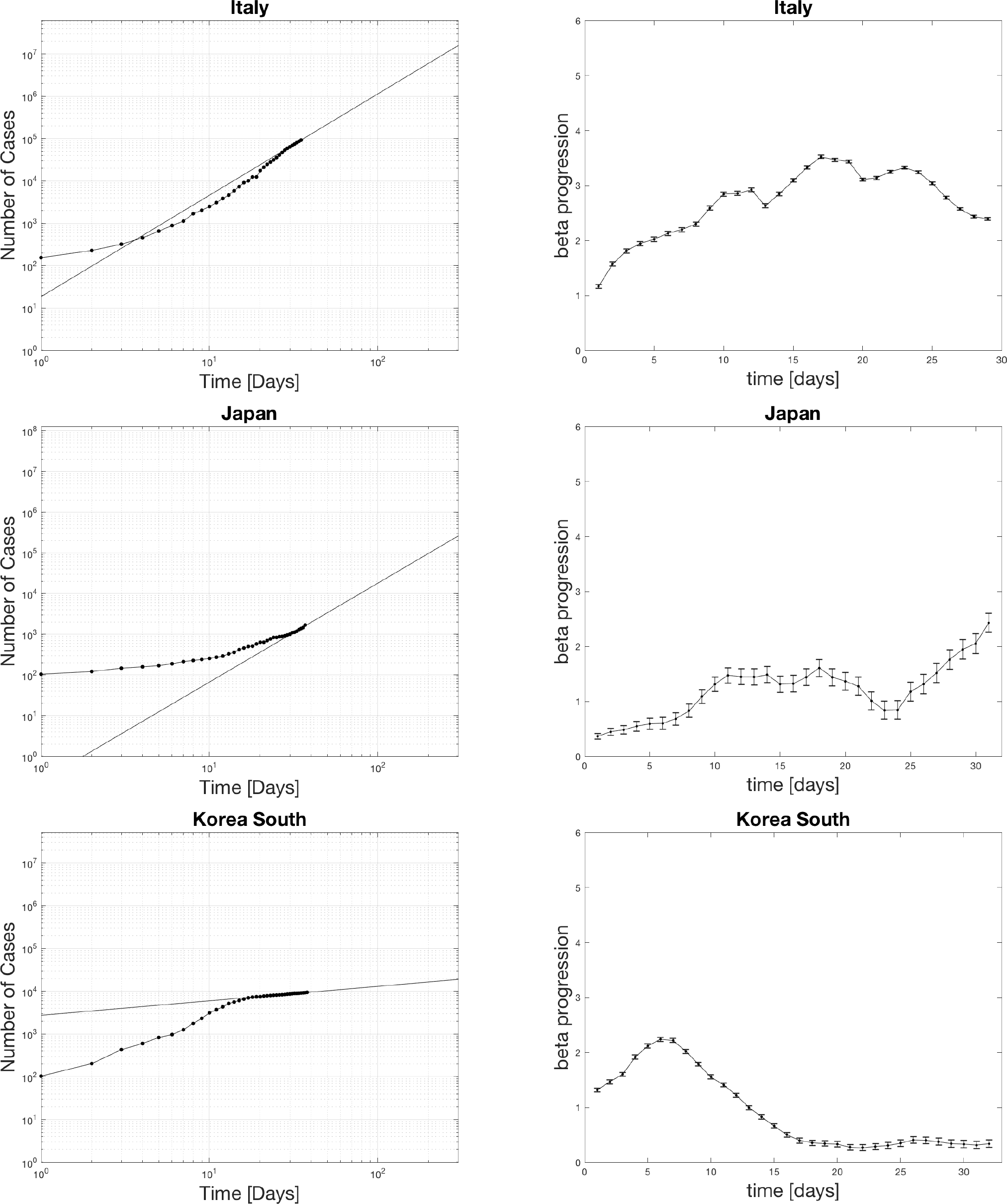

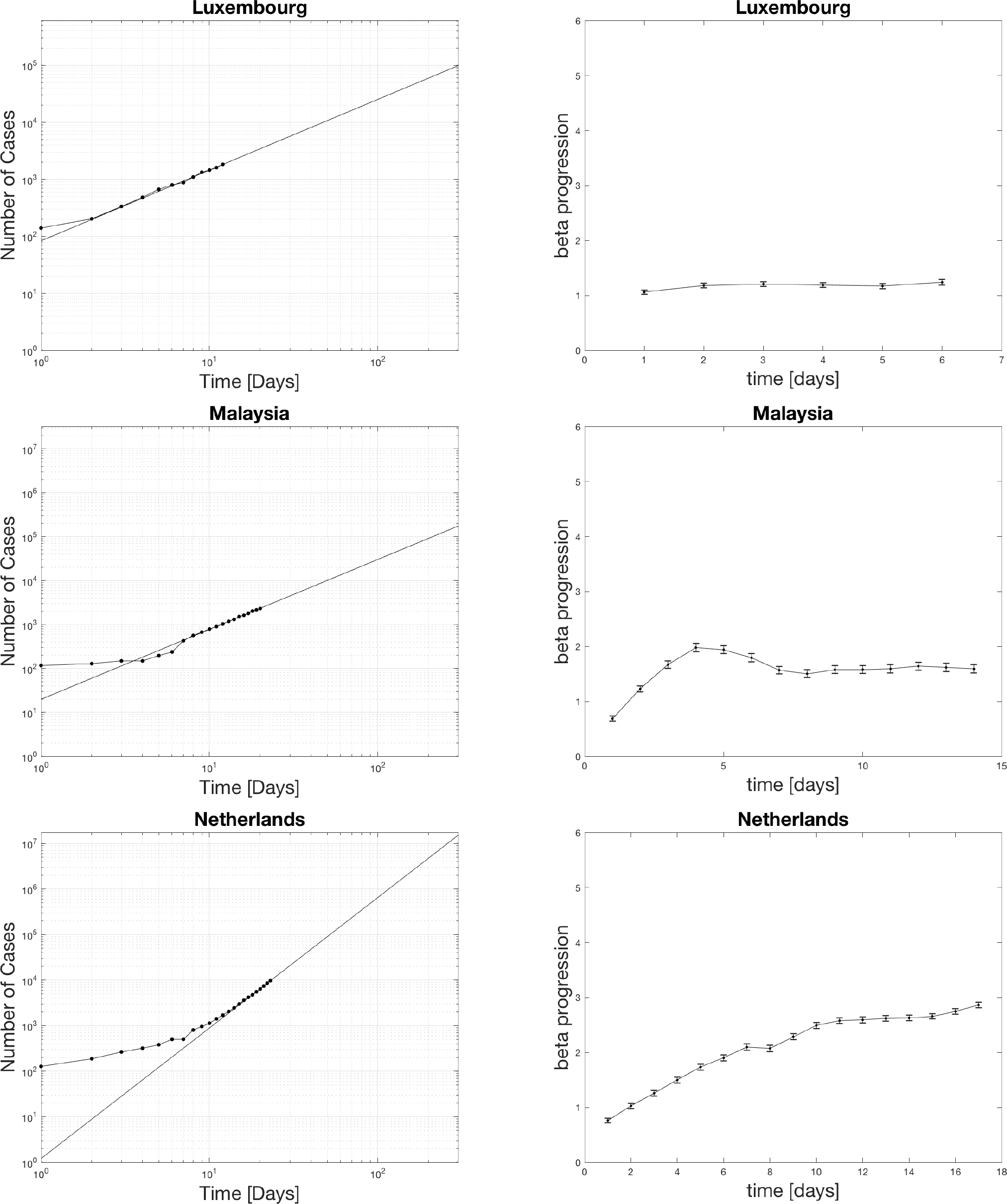

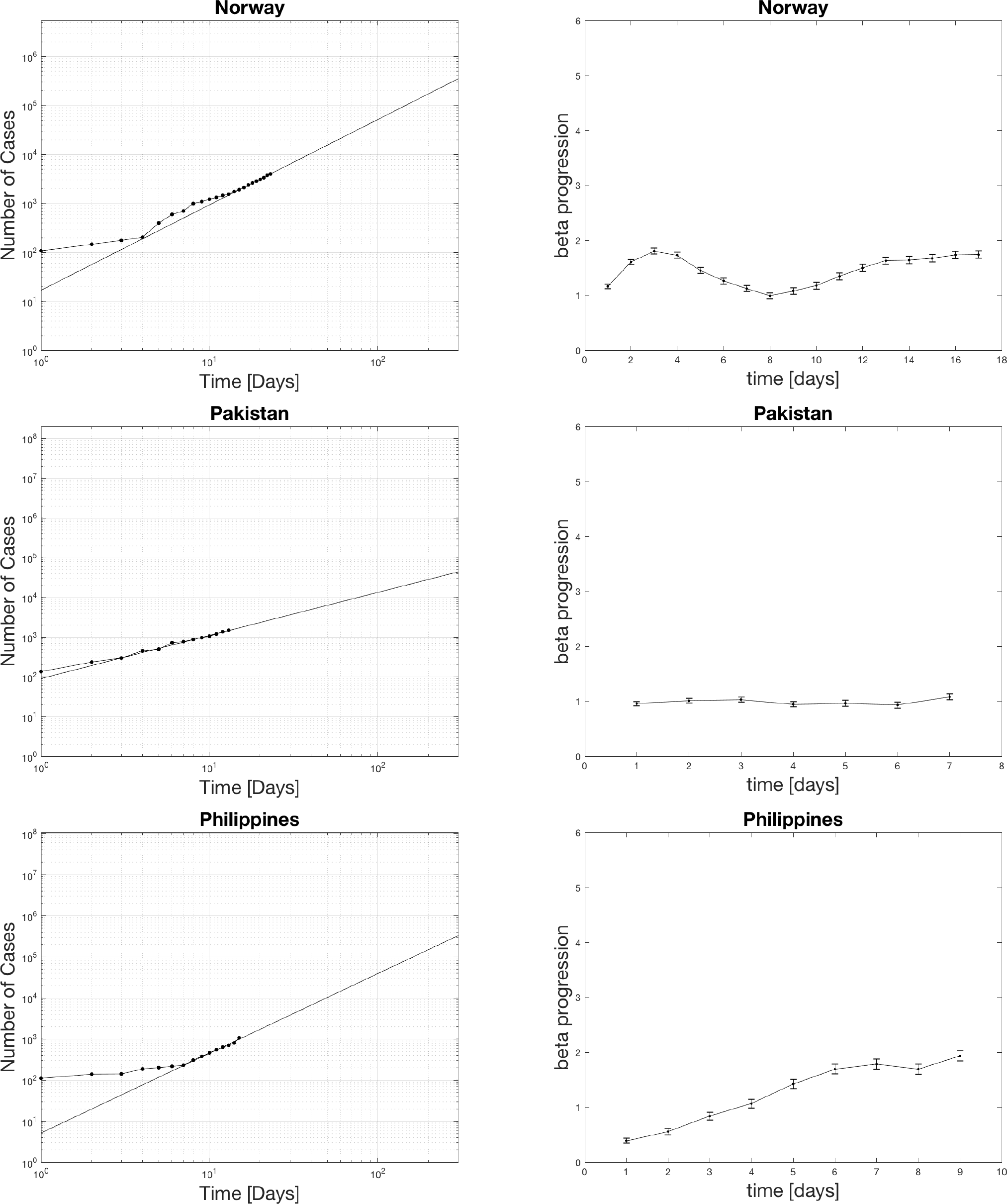

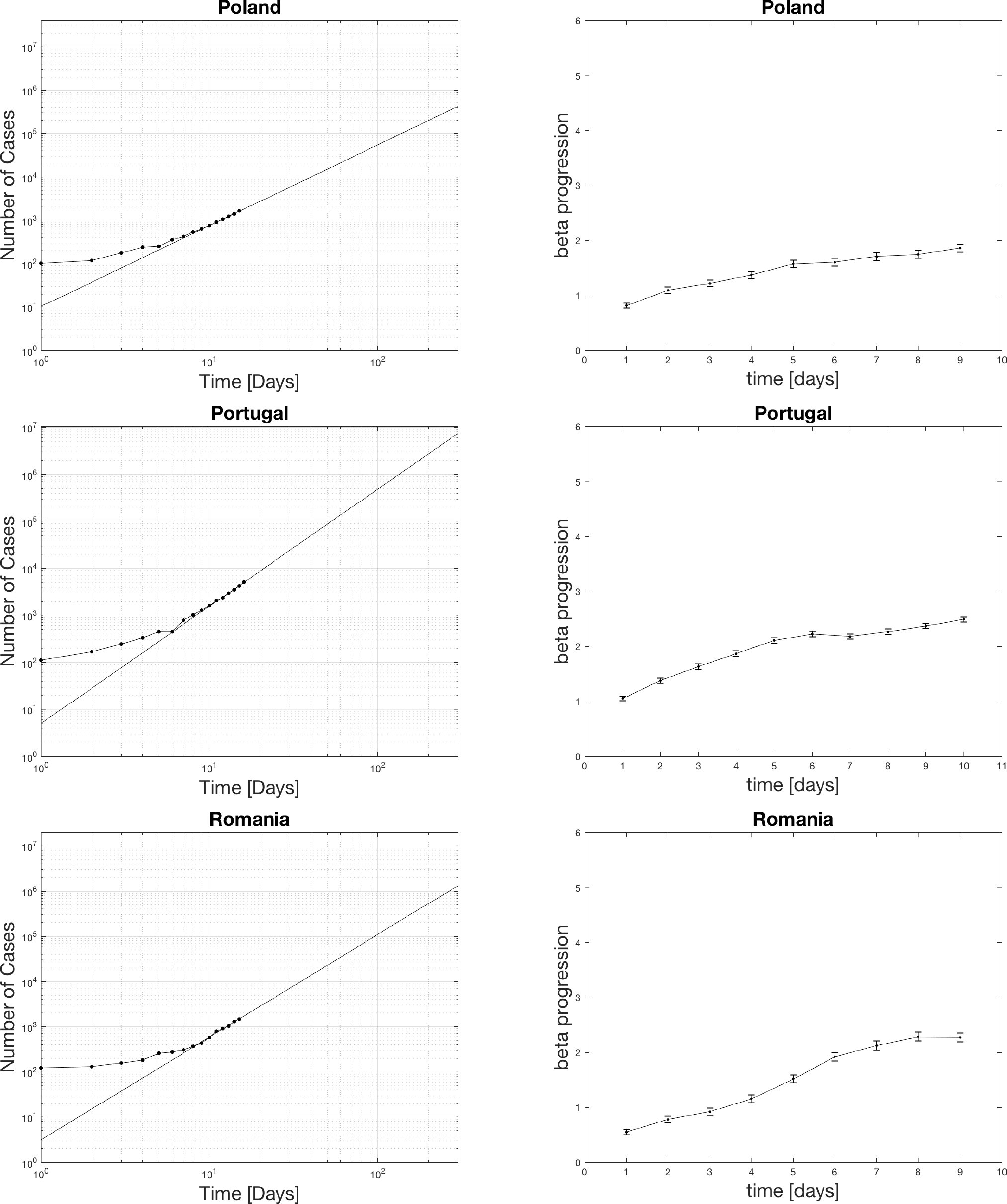

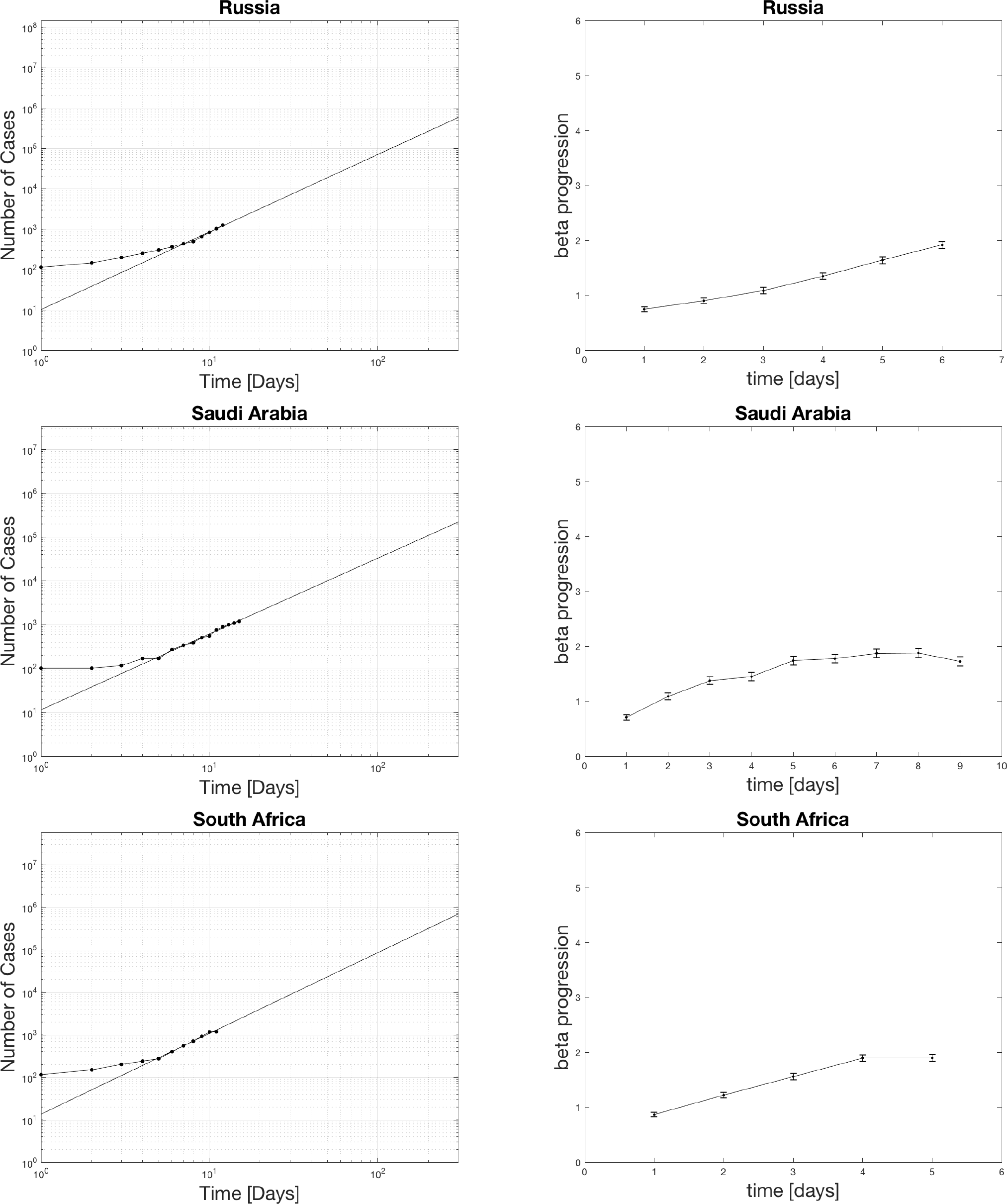

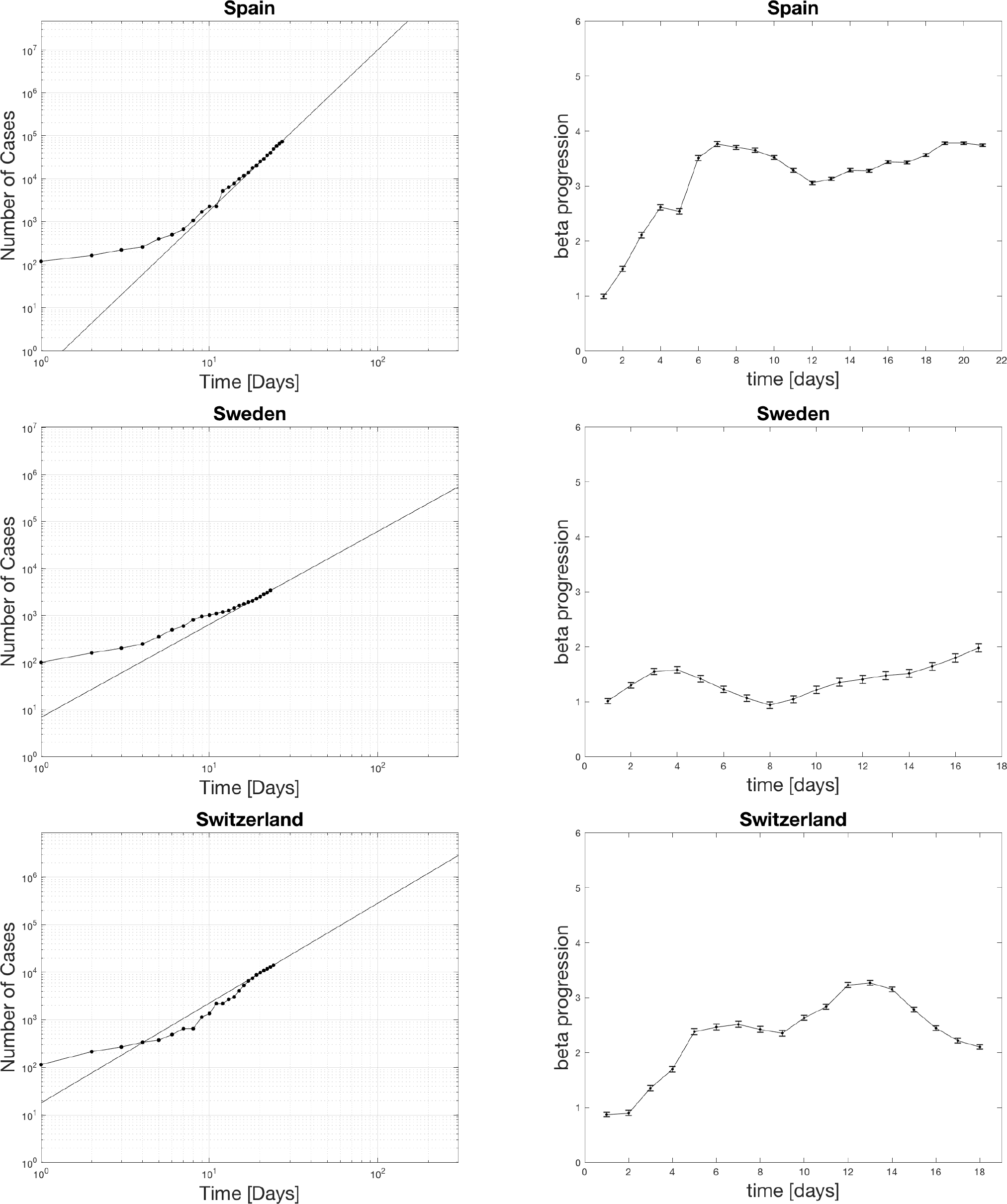

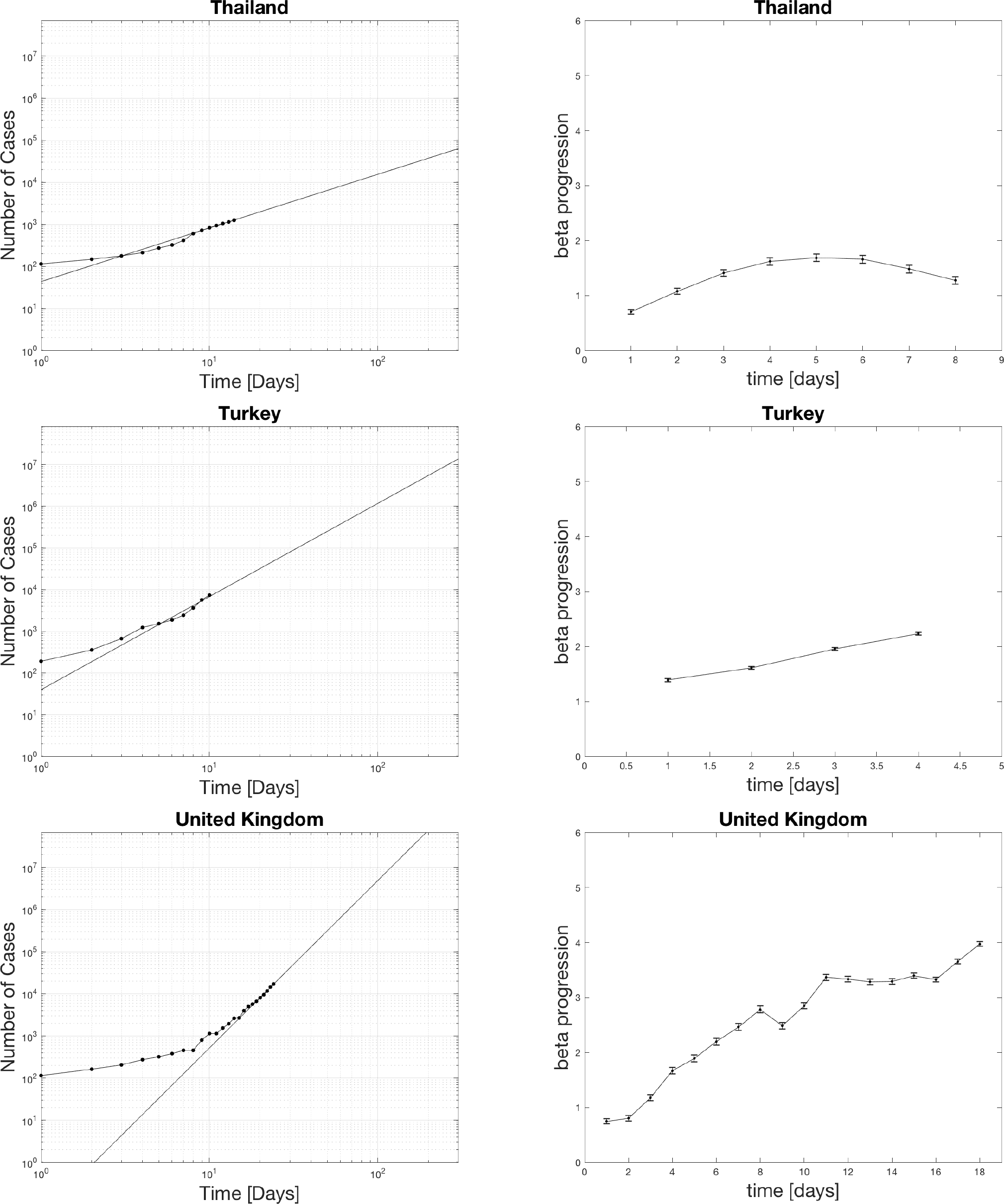

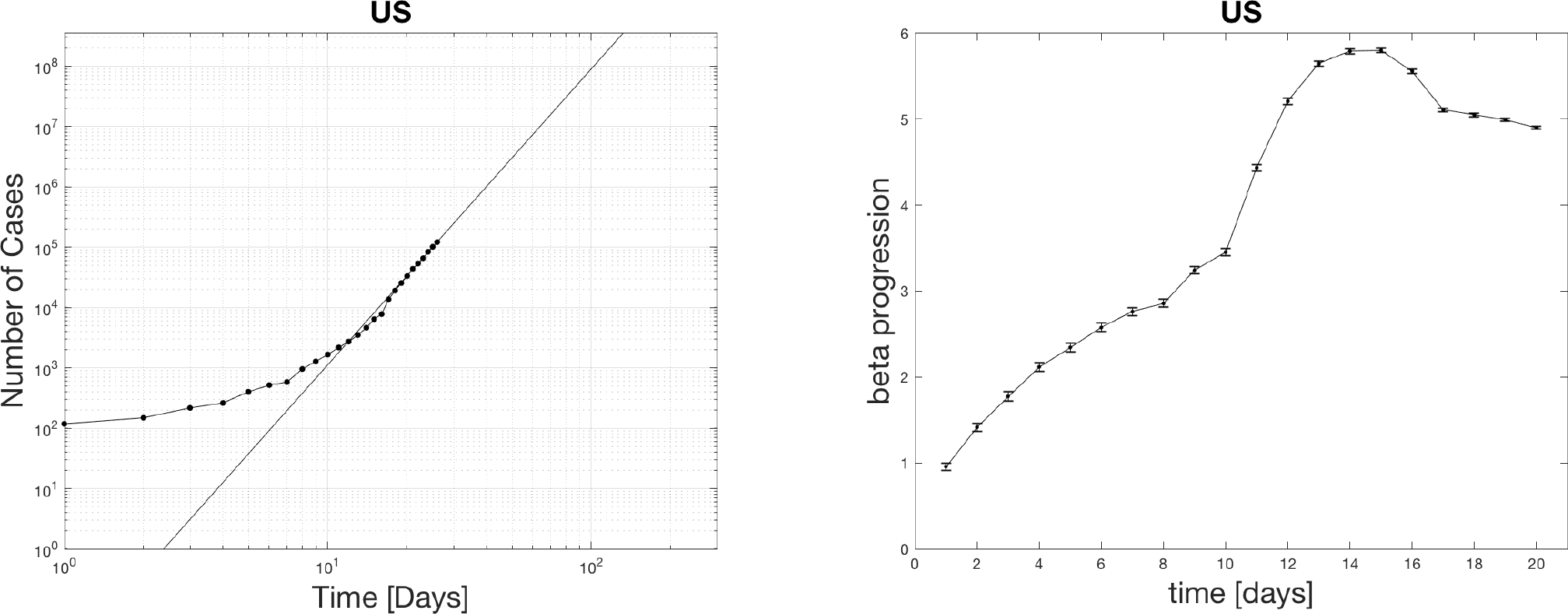

### 4.2 Distribution of beta currently in hot spots

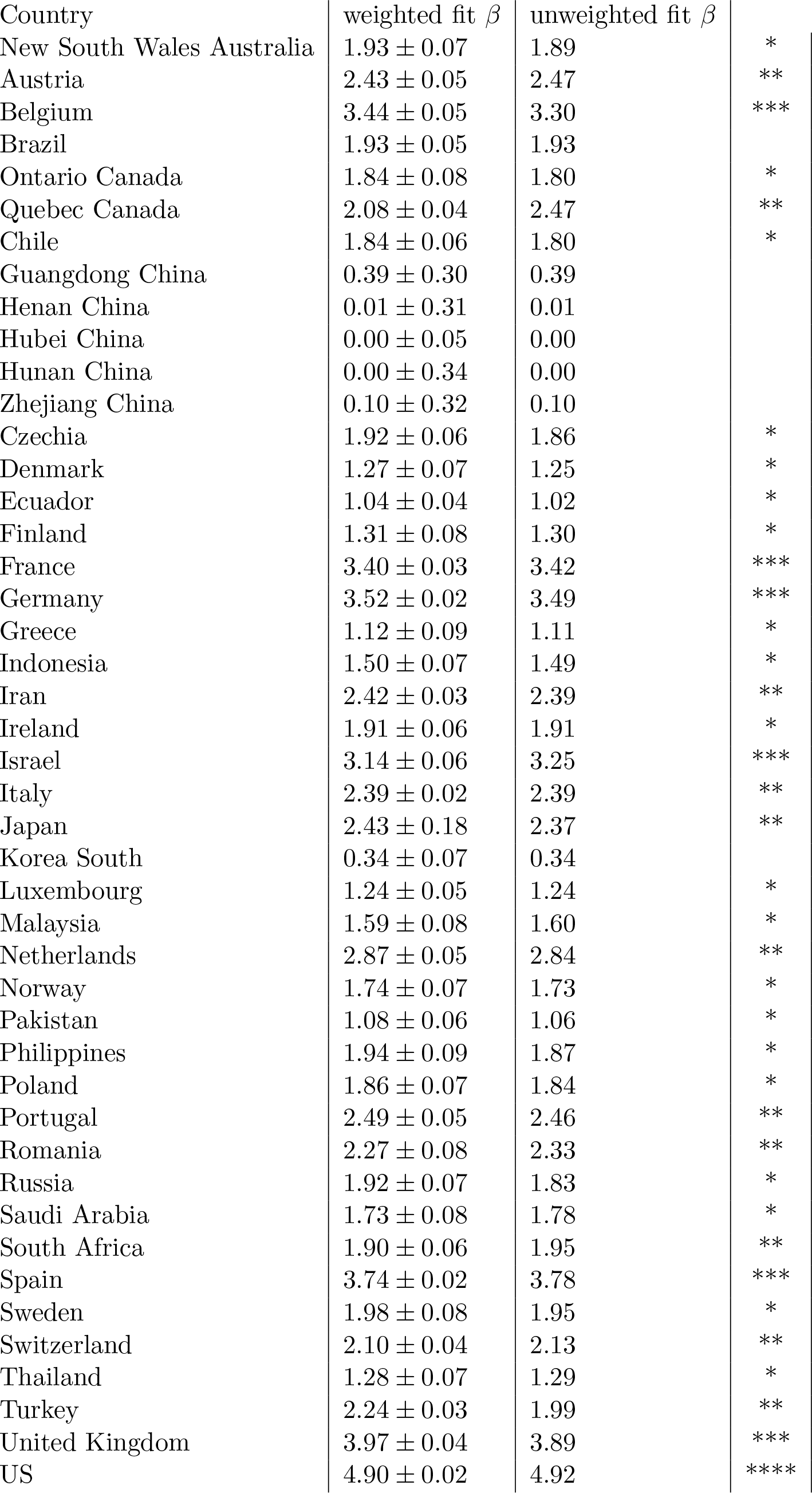

The countries with beta in the 1-2 range with one star, 2-3 ranges with two stars, 3-4 range with three stars, and 4-5 range with four stars. Beta picks out the most hard-hit countries such as the US, UK, France, Belgium, Germany, Spain, and Israel. Surprisingly Italy is only 2 stars now. The reason why Italy is doing so bad could be that they spent many more days at a middle range beta than some of the other countries who started later above 1000 cases. The US is the only country with four stars. It even crossed 5 at one point which is definitely significant. Many look closely to follow what happens in the US as this is the epicenter with the largest population.

Different software and algorithms would computer beta differently. There is not much difference if you try to find beta from a weighted curve fitting scheme where uncertainties are estimated from the square root of the the number of positive tested cases versus an unweighted curve fitting scheme. Doing an unweighted fit does not provide a direct way to find the measured error bar in beta versus the weighted method derived in the methods. The difference between the two methods is consistent with the actual computed error bars from the weighted analysis. The weighted curve fitting method from the methods is preferable.

## 5 Discussion

A plot of the 7-day window of power-law growth *β* or *D* is a good measure of the spread of infections in different populations. The dependence of *α* does not appear to indicate anything useful for the time being. The powers seem to vary between zero and just over five for the US, which is currently the epicenter. Perhaps this has to do with general behavior and connectedness of society rather when the disease first hit the country. The extrapolation of the power-law should enable us to make short term predictions of the number of cases in the future; however, social distancing measures can drop the power as indicated by measurements in South Korea and China. In those situations, the crossover occurs when *β* starts around 2 and takes three weeks or more from the point it starts to reduce. For the case of the US, this might take longer since the exponent is now near 5. log-log plots with the top scaled to the population of the country give some indication of how long a worst-case scenario could last before more or less everyone is infected. in the case, of the US this could be 100 days or less without mitigation. As we know however a power-law does not continue forever. This time scale should be compared to those for finding vaccines, supplying equipment, or finding medicine to treat COVID-19. There are several factors to consider in disease progression according to the number of positive cases and if this type of analysis relevant. Only people who are sick receive a test. Some people who are sick stay at home, and the disease resolves itself. Some people are positive but exhibit mild or no symptoms, so never receive a test. The data is only as reliable as countries continue to use the same set of testing standards throughout the pandemic. At some point, there could be so many cases that testing cannot keep up daily with the number of reagents required or available kits. After some point, it would be wise to implement serological testing or random testing of the general population to see a more accurate estimation of the total positive cases.

There are several factors to consider in disease progression according to the number of deaths. The numbers of deaths are time delayed from the date of the first infection. The time scale for death is longer than to become symptomatic. The number of deaths may increase in the coming days as hospitals become more overburdened, as is currently seen in Spain and Italy. There are fewer deaths than positive cases, so the sample sizes are smaller to determine the statistics. Whether by using positive cases or death rates, scientists should keep all their options open in tracking the course of COVID-19.

Many countries are mobilizing their manufacturing infrastructure to make additional personal protective equipment (PPE), respirators, and ventilators. One technique is to split the tubing of a ventilator to multiple patients. The timescale to produce this equipment should clearly be tens of days and not a hundred. The DIY community is also mobilizing to make 3D printed medical equipment rapidly. Companies like Dyson have created a ventilator in less than 10 days and plan to deliver 15,000 units. It is not clear if these types of numbers are enough to meet the actual future demand or if hundreds of thousands of ventilators or more are necessary. Presumably any person who died could have needed a ventilator. Spreading out the disease over time would allow more access to successful treatment.

Initiating social policy decisions in the initial stages of a pandemic is difficult, but we already saw how things unfolded in China with time to prepare. Unfortunately, most countries did little and now suffer the consequences. If the question is when is the best time to enact social distancing policies, the answer is now. The best time to plant a tree to get some shade is now if you did not do it twenty years ago. It is too soon to predict the final outcome in terms of infections and deaths by this method. Tracking a decrease in a beta down to 0-0.5 allows one to see the spread being mitigated. It is prudent to follow the examples of South Korea and China because they have demonstrated that COVID-19 can be stopped with sufficient effort. Sufficient effort in these cases meant extensive PCR testing, individual temperature checks at checkpoints, social distancing, limitations on gatherings, travel restrictions, quarantines, and wearing masks.

This analysis provides useful tools to see the progression of disease worldwide and whether conditions are getting better or worse. This analysis identifies the most hard-hit countries such as the US, France, Belgium, Germany, UK, Italy, Israel, and Spain with beta crossing over 3. South Korea and Chinese provinces have lowered their beta from 1.5-2.5 to near zero. Making optimistic predictions depends on what we do now. Countries besides South Korea and China are all in the middle phase. Let’s hope we can look towards lowering the beta in the affected area. This study is a step towards trying to visualize the progression of the disease over time to see if social policies are working or not. More automated visualizations [4] like these could help understand how to track the control of COVID-19 or the lack of it.

## Data Availability

The data file is available upon request.

## 6 Acknowledgements

Jack Merrin would especially like to thank Michael Sixt for encouraging me to think about these problems while working at home due to restrictions in place. I would like to thank Nick Barton, Simon Rella, Federico Sau, Ivan Prieto, and Pradeep Kumar for useful discussions.

P.S. Maybe it is time to stop shaking hands. Elbow bumps are not good either. Perhaps, we should just bow to each other. That seems civilized. Stay safe everyone.

